# A digital score of peri-epithelial lymphocytic activity predicts malignant transformation in oral epithelial dysplasia

**DOI:** 10.1101/2023.02.14.23285872

**Authors:** Raja Muhammad Saad Bashir, Adam J Shephard, Hanya Mahmood, Neda Azarmehr, Shan E Ahmed Raza, Syed Ali Khurram, Nasir M Rajpoot

## Abstract

Oral squamous cell carcinoma (OSCC) is amongst the most common cancers worldwide, with more than 377,000 new cases worldwide each year. OSCC prognosis remains poor, related to cancer presentation at a late stage indicating the need for early detection to improve patient prognosis. OSCC is often preceded by a premalignant state known as oral epithelial dysplasia (OED), which is diagnosed and graded using subjective histological criteria leading to variability and prognostic unreliability. In this work, we propose a deep learning approach for the development of prognostic models for malignant transformation and their association with clinical outcomes in histology whole slide images (WSIs) of OED tissue sections. We train a weakly supervised method on OED (n= 137) cases with transformation (n= 50) status and mean malignant transformation time of 6.51 years (±5.35 SD). Performing stratified 5-fold cross-validation achieves an average AUROC of ∼0.78 for predicting malignant transformations in OED. Hotspot analysis reveals various features from nuclei in the epithelium and peri-epithelial tissue to be significant prognostic factors for malignant transformation, including the count of peri-epithelial lymphocytes (PELs) (*p* < 0.05), epithelial layer nuclei count (NC) (*p* < 0.05) and basal layer NC (*p* < 0.05). Progression free survival using the Epithelial layer NC (*p* < 0.05, C-index = 0.73), Basal layer NC (*p* < 0.05, C-index = 0.70) and PEL count (*p* < 0.05, C-index = 0.73) shown association of these features with a high risk of malignant transformation. Our work shows the application of deep learning for prognostication and progression free survival (PFS) prediction of OED for the first time and has a significant potential to aid patient management. Further evaluation and testing on multi-centric data is required for validation and translation to clinical practice.

## Introduction

Oral cancer is amongst the most common cancers in the world and is considered a major health problem due to the significant associated morbidity and mortality ^1^. The 5-year survival rate has not improved over the last few decades regardless of improvements in surgical and oncological treatments. A large majority of oral cancers (>90%) are oral squamous cell carcinoma (OSCC) with one of the biggest obstacles to improvement in prognosis being delayed presentation of disease, as evidenced by the fact that survival for stage-I OSCC is 80% which reduces to 20-30% for stage IV disease ^2, 3^. OSCC is caused by a multitude of genetic and environmental factors and is preceded in a majority of cases by a potentially malignant state with proliferation of atypical epithelium known as oral epithelial dysplasia (OED) ^4^. Dysplastic lesions have been shown to have an increased risk of malignant transformation ^5^. Unfortunately, at present, there are no specific clinical tools, biological or molecular markers routinely used or recommended in clinical practice for prognostication of dysplastic lesions. Some clinical risk predictors have been suggested to be helpful including size, clinical site (e.g., floor of mouth, lower gums, lateral tongue), and clinical appearance (i.e. leukoplakia, erythroplakia etc.) and can be found in a wide range of conditions collectively referred to as oral potentially malignant disorders (OPMDs) in clinical practice ^6^.

In practice, OED diagnosis and grading are done on a tissue biopsy using histological assessment and light microscopy. The current gold standard grading system (the 2017 WHO grading system that uses three tiers of grading as mild, moderate or severe dysplasia) is subjective, taking into account at least 15 different cytological and architectural features as well as the extent of epithelial thickness involvement to determine the OED grade which guides treatment decisions ^4^. However, the cytological and architectural features are ill defined and lack in prognostic value e.g., mild or moderate OED can progress to malignancy while severe OED may not ^6^. In addition, OED grading suffers from significant inter- and intra-observer variation due to its subjective nature and interpretation can be hugely dependent upon the observer”s experience and training. To improve diagnostic reproducibility and prognostication, Kujan *et al*. ^7^ introduced the idea of a binary grading system, categorising cases as either low or high risk depending on the number of architectural and cytological features seen. Although reports have suggested improvement of diagnostic agreement and prognosis using the binary system, it also has shortcomings and has not been widely adapted for clinical use, highlighting the need for novel approaches ^6,8^ with objectivity and better prognostic value to inform patient management and aid treatment decisions ^9^.

Advances in the field of digital pathology and artificial intelligence (AI) have shown potential for improving histopathological diagnosis and prognosis, leveraging AI for objective and quantitative scoring of features. With wider adaptation of digital pathology in clinical practice, AI algorithms have also evolved and have shown promise in automated detection and quantification of histological features for classification ^10–14^, detection ^15–18^, segmentation ^19–21^ and survival analysis ^18,22^. Digitisation of histology slides generates digitised multi-gigapixel whole slide images (WSI), which can be used to develop algorithms to assist pathologists in diagnostic decision-making and better prognostication for improved patient management. To the best of our knowledge, there has been limited research on computational analysis of OED histology images for prediction of malignant transformation. Existing methods in the literature have used relatively small cohorts, manual elements, or region of interest (ROI) based analyses ^14,23–30^. All these methods have focused mainly on OED identification or grading and lack predictive or prognostic ability. Limited computational pathology work has been reported at the WSI level for predictive analysis of OED including recurrence and malignant transformation potential. There has also been variation in findings from studies reporting the correlation between OED grade, clinical and histological features, and malignant transformation. Dost *et al*. ^30^ examined 368 OED patients where 7.1% progressed to carcinoma and showed that there was no association of OED grade with malignant transformation. Gilvetti *et al*. ^31^ reported a study including 120 patients with a mean follow-up of 47.7 months (±29.9 SD) and showed that recurrence rate was significant in patients with erythroplakia with p = 0.023 with a mean time to recurrence of 62 months (±31.5 SD). Malignant transformation was also shown to have significant association with age (p = 0.034), clinical appearance (p = 0.030), lesion site (p = 0.007) and some other clinical features with a mean transformation time of 50 months (±32.5 SD). A recent study by Mahmood *et al*. ^32^ examined the correlation between individual histological features and OED prognosis. They examined OED biopsies from 108 patients with a minimum of five-year follow-up to analyse histological features predictive of recurrence and malignant transformation. Two different prognostic models based on presence of specific histological features (bulbous rete processes, hyperchromatism, loss of epithelial cohesion, loss of stratification, suprabasal mitoses and nuclear pleomorphism-irrespective of grade) were proposed with an area under the receiver-operator characteristic curve (AUROC) value of 0.77 for malignant transformation and 0.72 for recurrence. This highlights the usefulness of individual (grade-independent) histological features for OED prognosis prediction. A significant proportion of OED lesions can transform into malignancy (OSCC) and at present there are no tools available for objective and reproducible prediction of malignant transformation. Early prediction of malignant transformation is, therefore, crucial to aid patient care and inform appropriate treatment to improve prognosis and reducing the need for radical and disfiguring surgery at a later date.

In this study, we investigate the effectiveness of deep learning algorithms and nuclear features for prognostication from digitised WSIs of routine Haematoxylin & Eosin (H&E) stained OED histology sections in an end-to-end manner.

## Materials and Methods

### Data

The dataset used for this study comprised 163 Haematoxylin and Eosin (H&E) stained and scanned whole slide images (WSIs) of OED cases between 2005 to 2016. WSIs were scanned at ×20 using an Aperio CS2 scanner (n = 66) and at ×40 using a Hamamatsu scanner (n = 97) after ethical approval (REC Reference-18/WM/0335, NHS Health Research Authority West Midlands). Amongst 163 cases, 137 were OED cases and 50 cases which had transformed into malignancy. The remaining cases were non-dysplastic oral mucosal biopsies including benign hyperkeratosis or mild epithelial hyperplasia. The mean average age in the dataset of OED cases was 64.64 (range 25-97) with mean age for men (n = 84) was ∼66.3 and the mean age of women (n = 53) was ∼64.5. The main clinical sites of involvement were the tongue, floor of mouth and buccal mucosa. The mean time for malignant transformation was 6.51 years (±5.35 SD). The inclusion criteria for WSIs were decided upon the following conditions:

- A histological diagnosis of OED
- Sufficient availability of tissue
- Minimum five-year of follow-up data (including treatment, recurrence and transformation information) from the initial diagnosis
- Review of the pathology by two independent pathologists

More information about the cohort can be as seen in Table 1. Epithelium masks were obtained using HoVer-Net+ ^9^ and then refined manually for some cases, whereas slide-level labels were obtained for each case from patient records (i.e., clinical notes and biopsies) including histological grades, recurrence status, and malignant transformation status (i.e., OED has progressed into OSCC at the same diagnosed location within the follow-up time). The WSIs were split into train and test sets using 3 different stratified 5-folds on transformation status for all experiments. Patches of size 512 × 512 were extracted using the epithelium mask with an overlap of 50% from all the WSIs at 0.50µ per pixel (mpp). For extracting the deep features, ResNet-50 ^33^ was used as a feature extractor pre-trained on ImageNet. A feature vector of size 1024 was extracted for each patch resulting in a bag of shape *x* ∈ ℝ^*n*×1024^ for all WSIs (where *n* is the number of patches extracted).

**Table 1.**
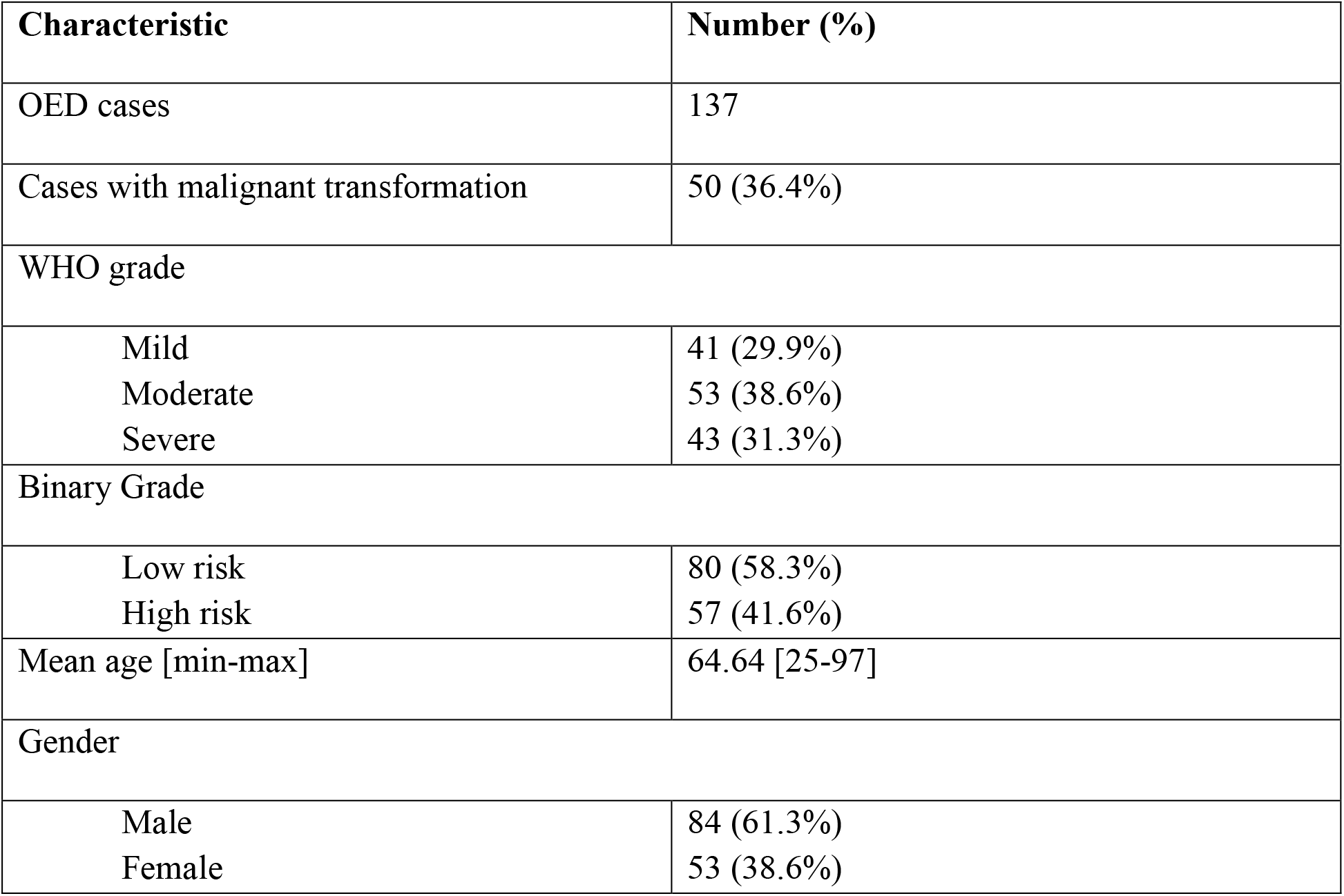
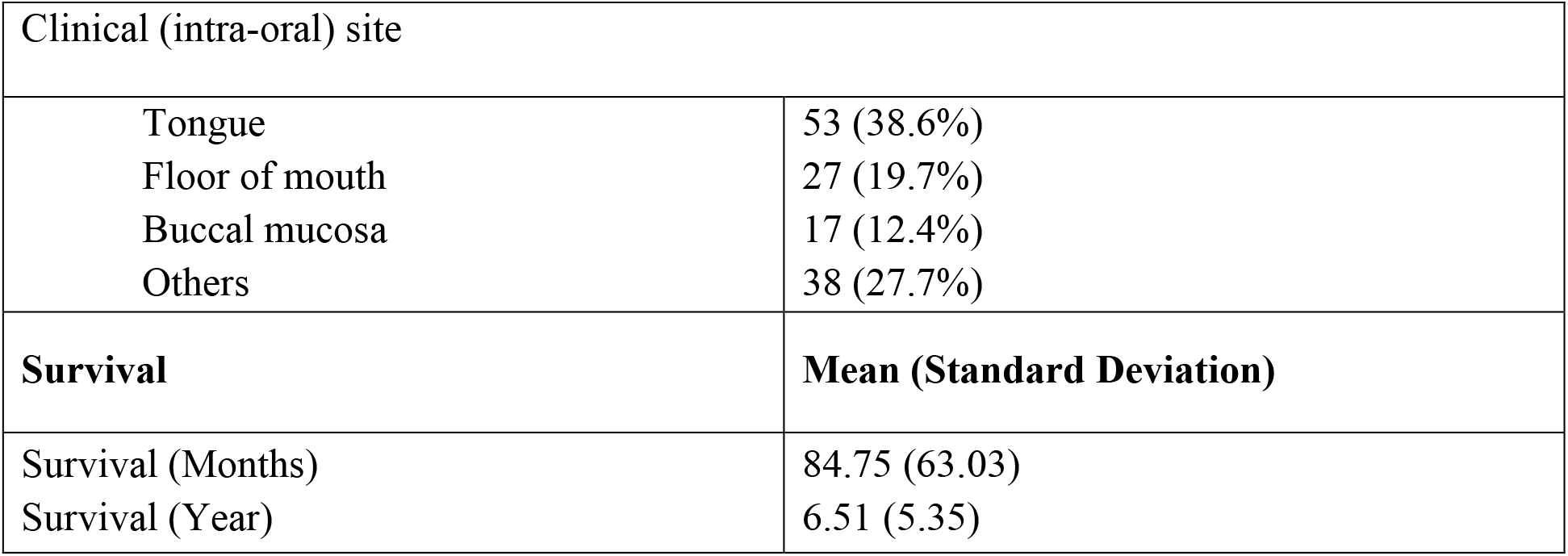
Characteristic of the cohort used for the study with clinical and demographic information of OED cases.

### Malignant Transformation Prediction

Figure 1 shows the overall pipeline, which involves initially extracting *X*patches of size *M*×*N*with slide level labels *Y*from WSIs with an overlap of *O*using the epithelium mask. Extracted patches were utilised for training the deep learning models for predicting malignant transformation. In this study, we used iterative draw-and-rank sampling (IDaRS) ^34^ which works by ranking and selecting the top and random patches from a WSI assuming that not all patches are equally important and predictive of the outcome. IDaRS selects two subsets of patches for training including random patches *r* and top-ranked patches *k* for each WSI. Both subsets are then pre-processed using the standard set of augmentations and train a CNN with weak labels. We have also compared the IDaRS with other fully supervised and weakly supervised algorithms e.g., multi-layer perceptron (MLP), Attention-MIL (A-MIL) ^35^, clustering constrained attention multiple instance leaning (CLAM) ^36^, and CNN based benchmark classification models (ResNet ^33^, DenseNet ^37^ and Vision Transformers ^38^ with max pooling as an aggregator for the final WSI label).

**Figure 1.**
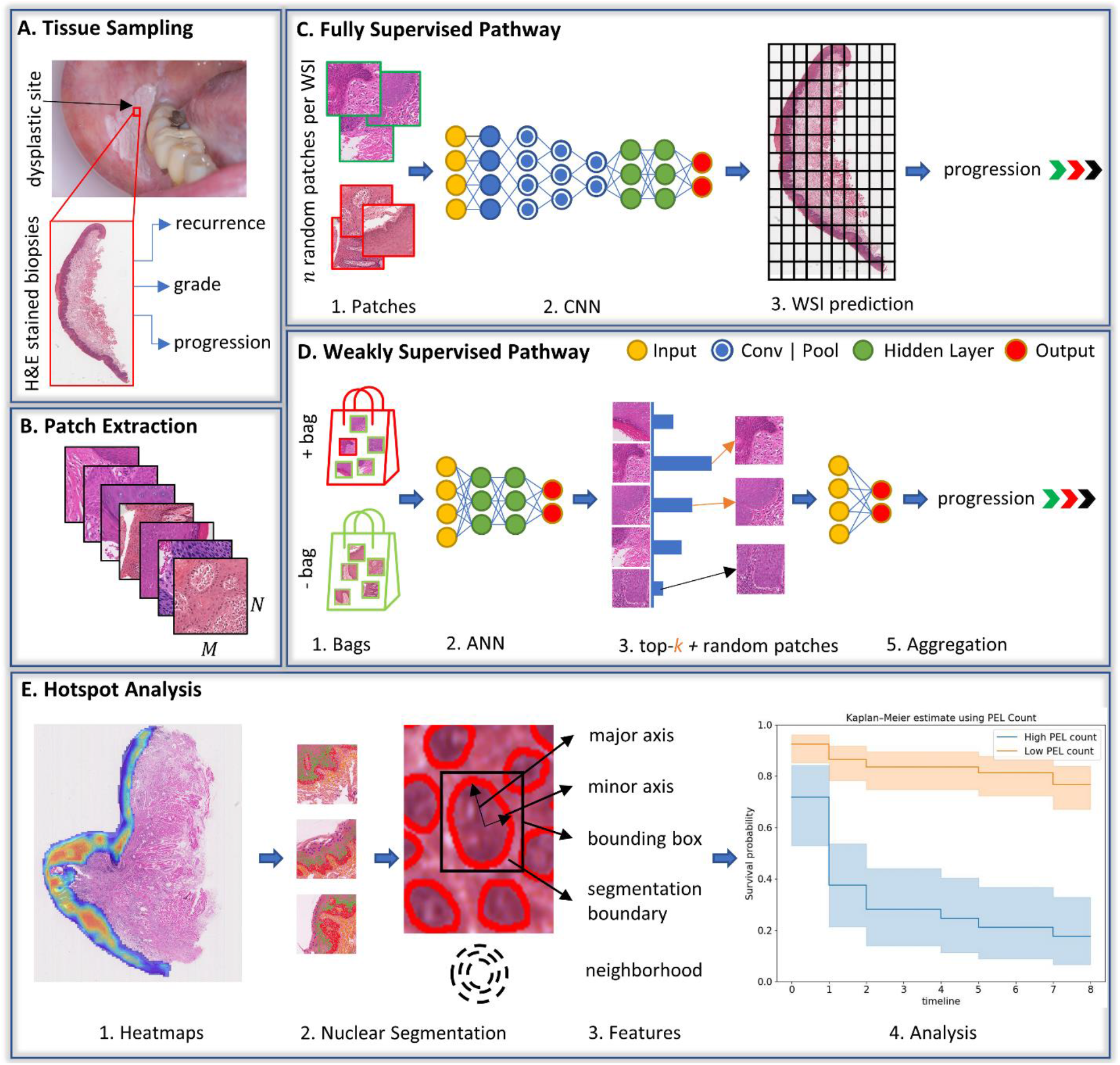
Overall workflow of the study is shown in different sections. A) the process of getting the tissue biopsies from dysplastic lesions and corresponding WSIs with their associated labels assigned by a pathologist. B) patches of size M×N were extracted from the epithelium region of WSIs. C) fully supervised pipeline where the patches were assigned the WSI level labels and trained using CNNs for the downstream tasks. D) weakly supervised pipeline where positive (+tive) and negative (-tive) batch of features/images was created and used for training. E) heatmaps were generated using IDaRS to explore the hotspot areas and their contribution towards the malignant transformation prediction using nuclear analysis. Nuclear features from the hotspot and cold spots were used for progression free survival.

### Cellular Composition Analysis

To further analyse and validate the hotspots being identified by the IDaRS model cellular compositions of top tiles (i.e., hotspots and coldspots) from transformed and non-transformed cases were analysed. Nuclear features were extracted from each layer (i.e., keratin, epithelial, basal) and associated connective tissue in an automated manner using the nuclear segmentation and classification. For this purpose, input patches were first stain normalised using a sample from The Cancer Genome Atlas (TCGA) cohort before being fed into HoVer-Net ^15^ which was pre-trained on the PanNuke dataset ^21^ for nuclear instance segmentation and classification. For segmentation of the keratin, epithelial and basal layers within the epithelium, HoVer-Net+ ^9^ was used. Table 2 shows a range of morphological and proximity features extracted from the segmented image patches and aggregated statistically using the minimum ∧, maximum ∨, mean μ, median *m* and standard deviation σ. Here, ordinary least square (OLS) was used with post-hoc t-tests for calculating the statistical significance with Benjamini/Hochberg adjustment ^39^. Cellular composition helps understanding/interpreting the results of IDaRS and differentiate transformed cases from non-transformed ones in an objective manner.

**Table 2.**
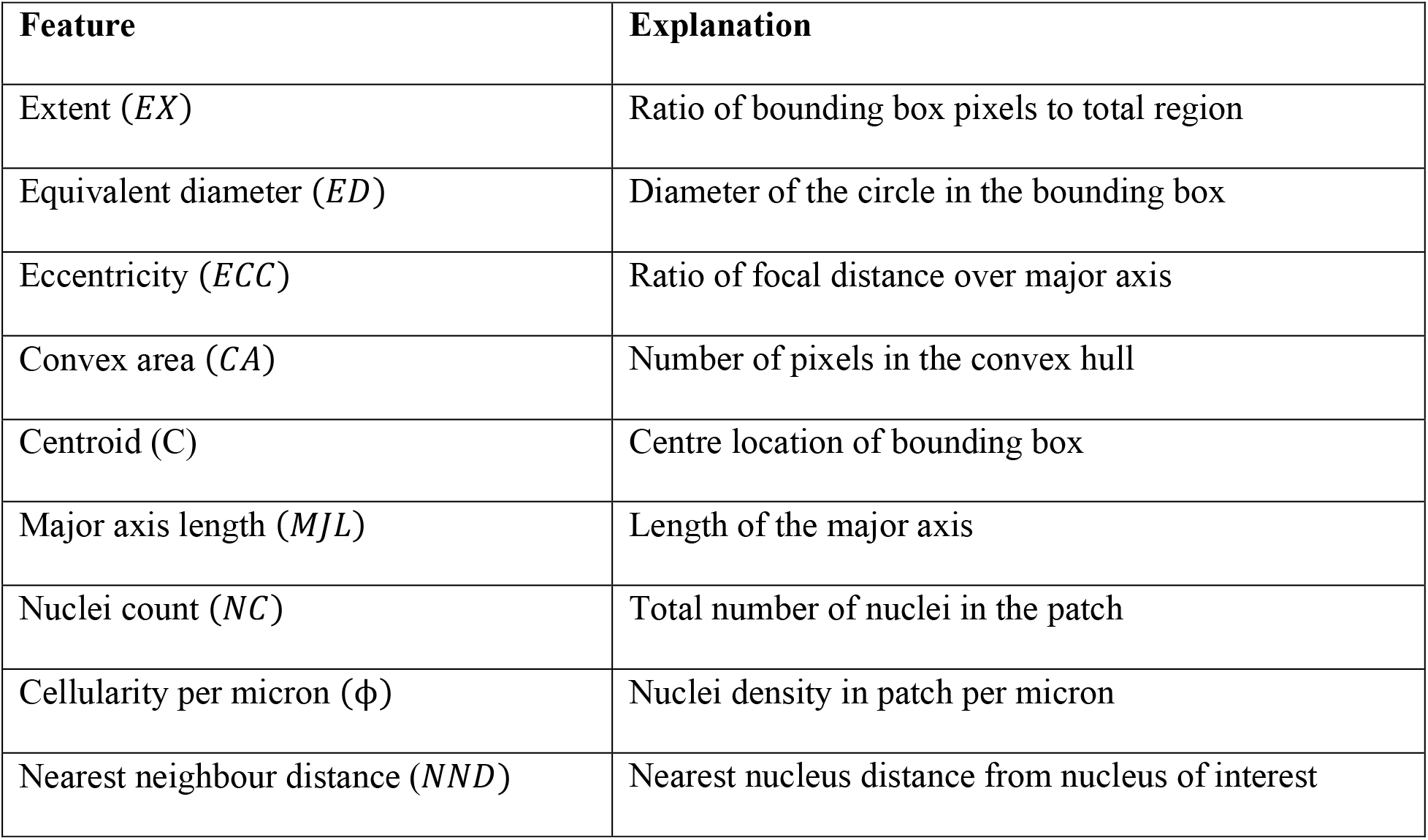
Nuclear features extracted from layer wise nuclei and their explanations.

### Peri-epithelial Lymphocytes (PELs) Count

Elevated PEL counts can be linked to a higher risk of malignant transformation in oral epithelial dysplasia (OED) and in order to further explore the role of PEL count in transformed and non-transformed cases, a Wilcoxon rank-sum test was performed where *p* < 0.05 was considered significant. Moreover, we also analysed the distributions of PEL count in subgroups based on two clinical features i.e., gender and age. Gender was divided into male and female groups, the age subgroups were separated into ranges between 0-50, 51-70, and 71-100.

### Survival Analysis

To investigate the prognostic significance of the clinical, pathological, and nuclear features for progression free survival (PFS), Kaplan–Meier (KM) curves and Cox proportional hazard (CPH) model were used for univariate and multivariate analysis. To distinguish between the high risk (short term survival) and low risk (long term survival) groups the optimal cut off value was calculated by taking the mean of hazard value for each instance using CPH model where the statistical significance is large between the high and low risk groups. Furthermore, a long-rank test was performed to determine the statistical significance and *p* < 0.05 was considered statistically significant.

### Experiments

For IDaRS, we set the random patches *r* = 30, top-patches *k* = 5 and trained a pretrained ResNet-34 on ImageNet with a batch size of 16 and patch size of 256. IDaRS is trained for 30 epochs with binary cross-entropy loss and optimised using the Adam optimiser. For training, MLP and CLAM deep features were then fed as an input to the models for generating WSI-level outputs. MLP and CLAM were trained for 1000 epochs using the default configurations from the CLAM ^36^. For A-MIL and CNN models” the same input and configurations as IDaRS were used for the training and test purposes. All models were trained and tested on a system with two Nvidia Titan-X with 12 GB of memory, dedicated RAM of 128GB, and an Intel® Core i9 processor.

To validate the results, stratified on transformation status 5-fold cross validation was performed three times with different random seeds. AUROC and F1-score (macro) were used as performance metric and are averaged across the folds. F1-score can be though as weighted mean between the precision and recall known as harmonic mean, and is calculated as:

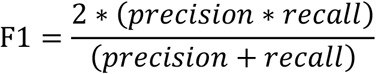

F1-score (macro) computes the arithmetic mean of F1-score per class treating all classes equally and regardless of their number. AUROC evaluates the binary problems by plotting true positive rate (TPR) against the false positive rate (FPR) at various thresholds. The area under the ROC curve (AUROC) measures the ability of the classifier to differentiate between the two classes where the TPR and FPR are calculated as:

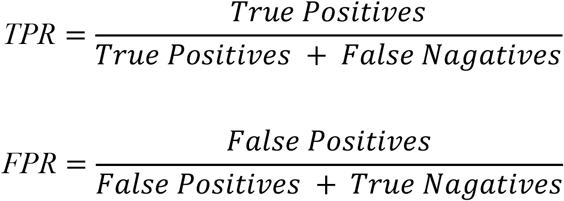

## Results

### Malignant Transformation

Our experiments shown in Table 3 indicate that the performance of IDaRS is comparatively better than the other weakly and fully supervised algorithms with an AUROC of 0.78 (±0.07 SD) and F1-score of 0.69 (±0.05 SD) as compared to MLP, CLAM, and A-MIL. It can also be observed from the ROC plots in Figure 2 that the standard deviation across different folds for IDaRS is smaller as compared to the other weakly supervised algorithms. It is worth noting that the performance of CLAM is competitive to IDaRS as compared to the MIL in terms of F1-score. The reason for poor performance of CLAM can be attributed to less confident predictions as a consequence to the fixed input feature representations leaving small room for optimal thresholding. The fully supervised networks performed worse compared to other weakly supervised models due to the inherent nature of the problem which introduces noise in the labels corrupting the model”s training.

**Table 3.** Performance of IDaRS model as compared to other weakly supervised and fully supervised models with deep features where IDaRS is achieving high performance in terms of AUROC.

| Model | top-k | AUC ± SD | F1-score ± SD |
| --- | --- | --- | --- |
| MLP | 1 | 0.65 ± 0.09 | 0.56 ± 0.11 |
| | 5 | 0.64 ± 0.11 | 0.55 ± 0.01 |
| Attention-MIL <sup>35</sup> | - | 0.54 ± 0.07 | 0.44 ± 0.03 |
| CLAM <sup>36</sup> | 1 | 0.65 ± 0.04 | 0.64 ± 0.04 |
| | 5 | 0.65 ± 0.05 | 0.63 ± 0.01 |
| IdaRS <sup>34</sup> | 5 | **0.78 ± 0.07** | **0.69 ± 0.05** |
| ResNet-50<sup>33</sup> | - | $0.54 \pm 0.10$ | $0.43 \pm 0.11$ |
| ViT<sup>38</sup> | - | $0.55 \pm 0.01$ | $0.45 \pm 0.08$ |
| DenseNet<sup>37</sup> | - | $0.56 \pm 0.05$ | $0.44 \pm 0.01$ |

**Figure 2.**
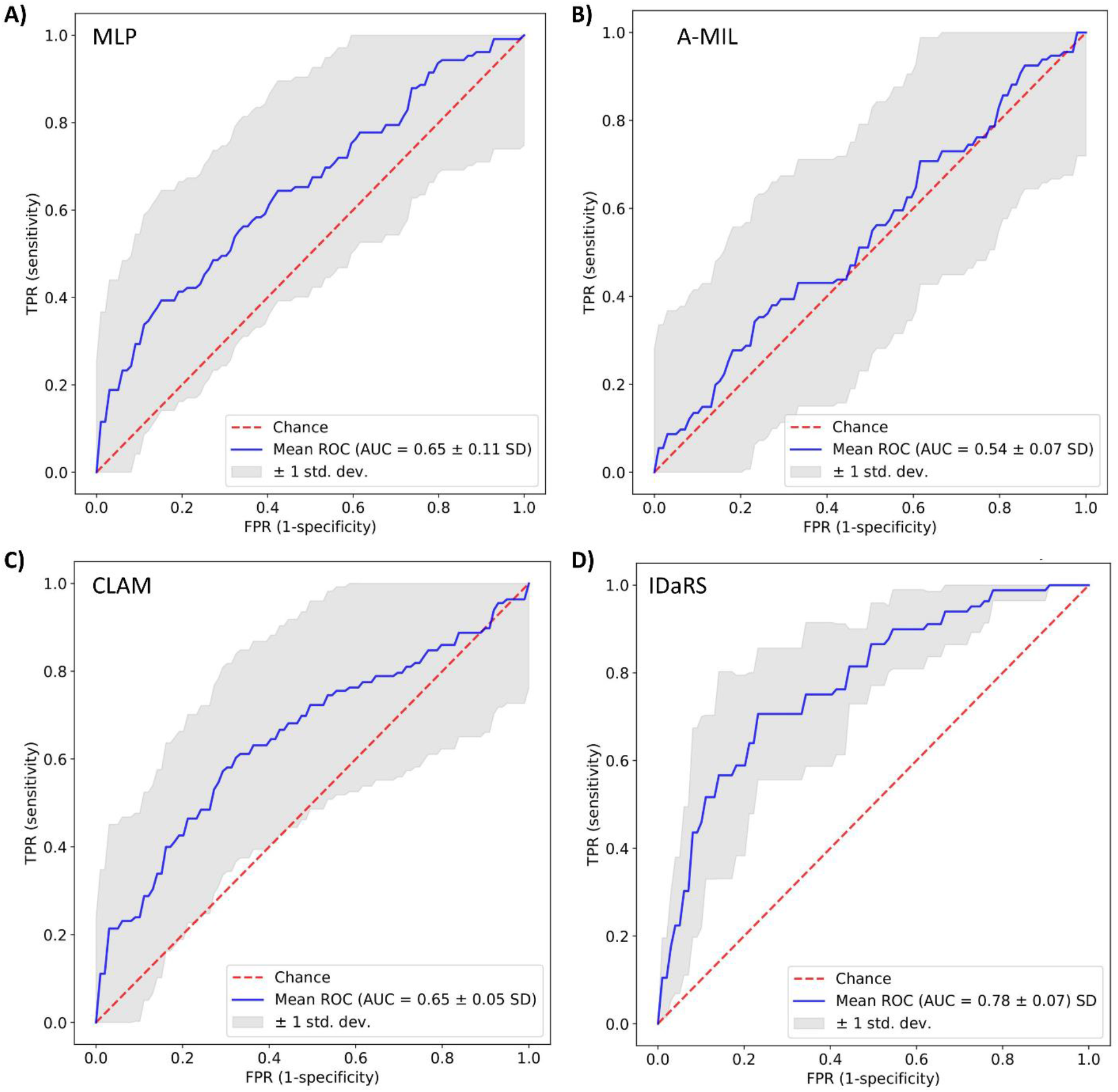
ROC curve plots on 5-fold cross validation for OED malignant transformation prediction using (A) MIL (B) A-MIL (C) CLAM and (D) IDaRS.

### Exploring the visual patterns

To validate and further investigate the features learnt by the top performing IDaRS, we explored the top tiles from the heatmaps of transformed and non-transformed WSIs. For correlating the hotspot/coldspots with the clinical features, heatmaps were also analysed manually for corroboration purposes by an expert pathologist SAK. Figure 3 shows the heatmap for a histologically high risk case where the red (hotspot) colour represents a region with higher probability of malignant transformation while the blue (coldspot) colour corresponds to a region with a low probability of transformation. Closer examination of hotpots shows evidence of disordered stratification, dyskeratosis as well as nuclear and cellular pleomorphism with a dense lymphocytic infiltrate in the adjacent peri-epithelial connective tissue. The dense lymphocytic infiltrate is referred to as peri-epithelial lymphocytes (PELs) for the rest of the analysis.

**Figure 3.**
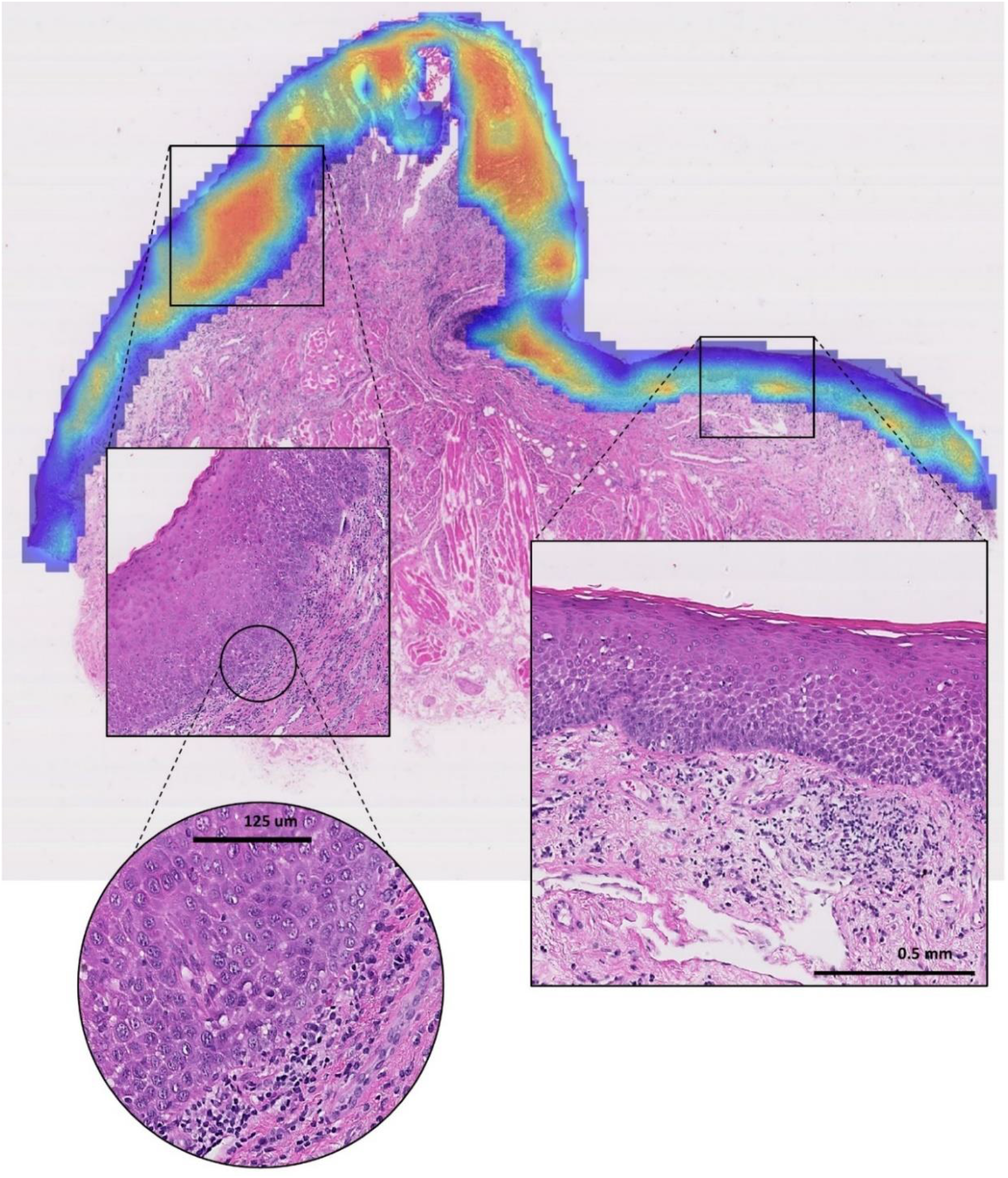
Heatmap for the malignant transformation predicting using IDaRS. The red region shows the high probability of malignant transformation in those areas.

### Cellular Composition Analysis

Following the manual analysis of the heatmaps, automated cellular composition analysis was performed to uncover significant hidden patterns/features in transformed vs non-transformed cases. Table 4 shows the prognostic significance of the extracted nuclear features for predicting malignant transformation. For epithelial layer, variation in eccentricity (*p* = 0.048), bounding box (*p* = 0.0487) and total nuclei count (*p* < 0.0001) showed significance along with basal layer NC (*p* < 0.0001). An increase in cell count (hyperplasia or crowding) is an important feature observed in high risk dysplasia in both the central epithelium layer and specifically within the basal layer. Other features in epithelium e.g., variation in nuclei count (100µ per pixel) and nearest nuclei distance corresponds congestion in spatial arrangements of epithelial nuclei and requires more data for validation. Similarly, changes in basal layer nuclei minor axis, equivalent diameter corresponds to the nuclear pleomorphism and are observed in high risk OED cases. Interestingly, the nuclei count in the connective tissue area also showed significance for predicting the transformation (*p* = 0.0004) which corresponds to the previous observation regarding the dense lymphocytic infiltrate in the adjacent peri-epithelial connective tissue.

**Table 4.**
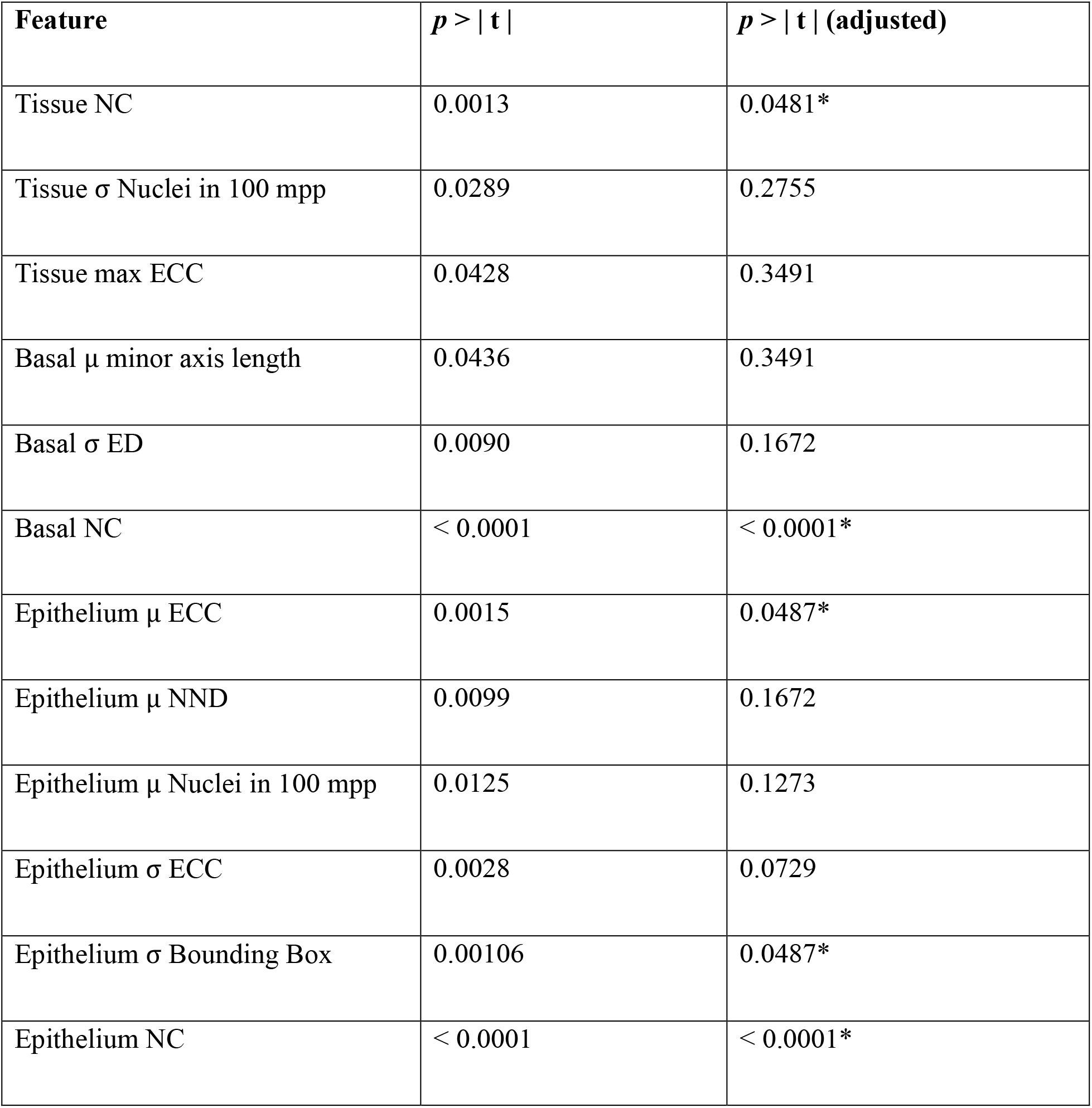
Ordinary least square regression for malignant transformation with t-test significance of nuclear features with Benjamini/Hochberg ^39^ adjustment. Significant p value is is highlighted using *.

| **Feature** | **$p > \vert t\vert $** | **$p > \vert t\vert $ (adjusted)** |
| --- | --- | --- |
| Tissue NC | 0.0013 | 0.0481* |
| Tissue $\sigma$ Nuclei in 100 mpp | 0.0289 | 0.2755 |
| Tissue max ECC | 0.0428 | 0.3491 |
| Basal $\mu$ minor axis length | 0.0436 | 0.3491 |
| Basal $\sigma$ ED | 0.0090 | 0.1672 |
| Basal NC | < 0.0001 | < 0.0001* |
| Epithelium $\mu$ ECC | 0.0015 | 0.0487* |
| Epithelium $\mu$ NND | 0.0099 | 0.1672 |
| Epithelium $\mu$ Nuclei in 100 mpp | 0.0125 | 0.1273 |
| Epithelium $\sigma$ ECC | 0.0028 | 0.0729 |
| Epithelium $\sigma$ Bounding Box | 0.00106 | 0.0487* |
| Epithelium NC | < 0.0001 | < 0.0001* |

### Peri-Epithelial Lymphocytes (PELs)

Figure 4 shows examples patches from both hotspots (red) and coldspots (blue) regions of the transformed and non-transformed cases with their corresponding layer-wise cellular compositions. For most of the coldspots, the epithelium and basal nuclei are dominant whereas in the hotspots (red) PELs are in abundance in the transformed cases compared to non-transformed cases. As a whole, PELs were statistically significant (*p* = 0.02) for differentiating between the transformed vs non-transformed cases. Gender based subgrouping showed no significance between male and female groups. However, for age, 0-50 group showed prognostic significance with respect to malignant transformation with *p* = 0.001. Figure 5 shows the boxen plots for (A) the overall distribution of PEL ratio in transformed cases versus non-transformed cases and (B) the distribution of PEL ratio in transformed cases versus non-transformed cases including age subgrouping.

**Figure 4.**
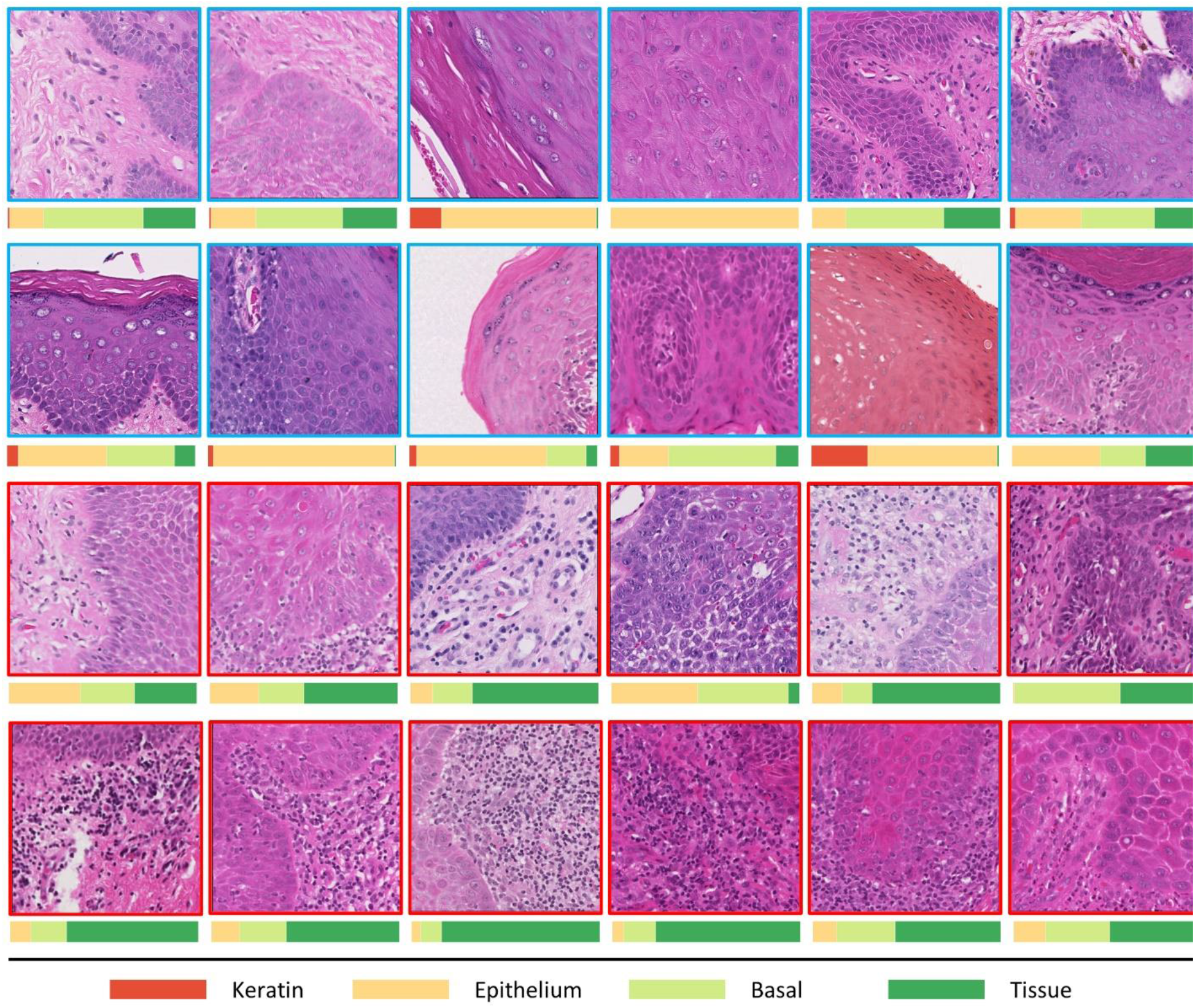
Patches extracted from the hotspot (red) and coldspots (blue) of the WSIs with their layer wise nuclear composition. Most of the coldspot regions have dominant epithelial nuclei as compared to the hotspots where PEL can be seen dominating the overall ratio.

**Figure 5.**
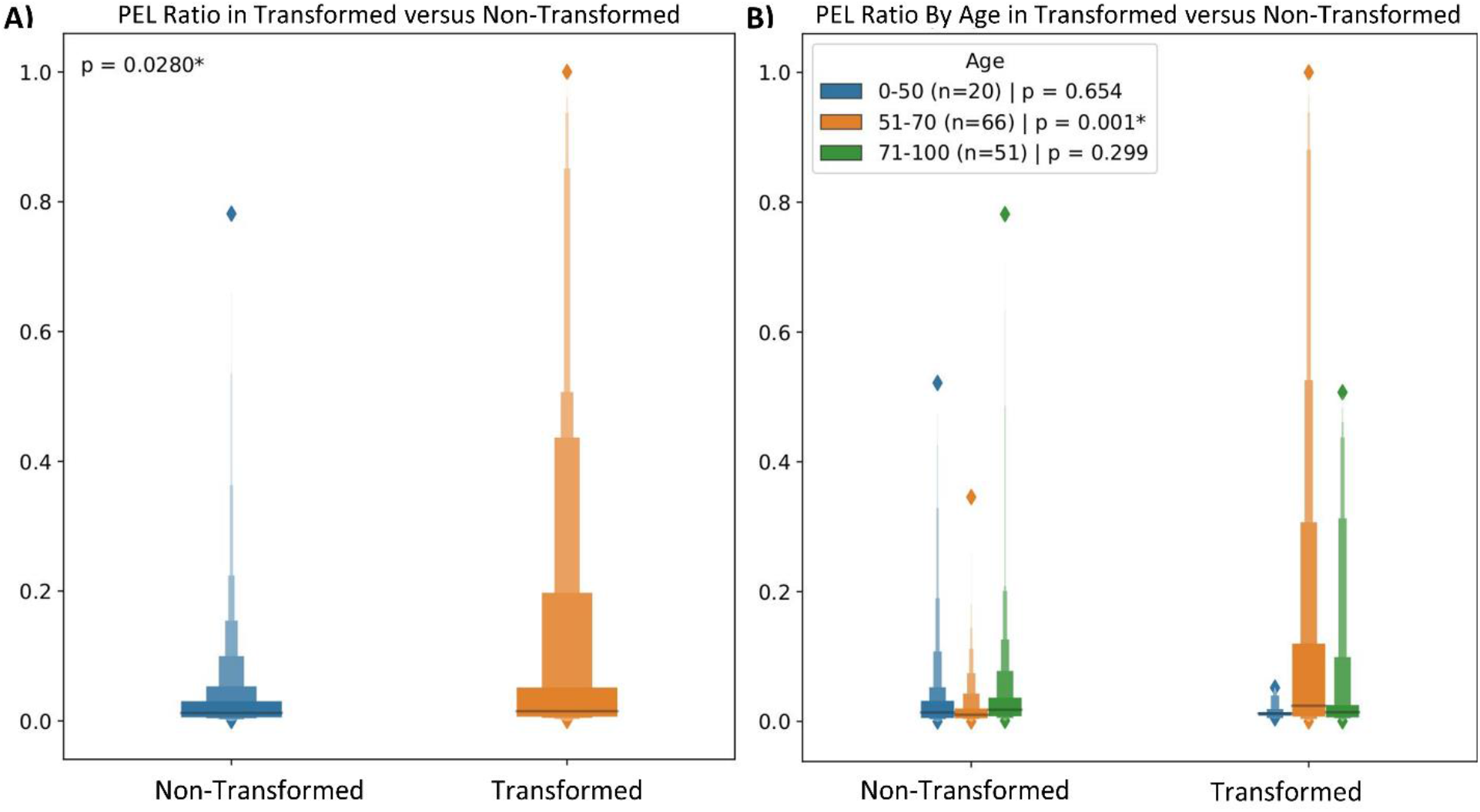
(left) Shows the boxen plot for the ratio of PELs present in both transformed and non-transformed patches and (Right) Shows the further breakdown of the PEL ratio in age groups where the 0-50 age group has distinct difference in PEL ratio as compared to the other groups.

### Survival Analysis

Table 5 shows the univariate analysis of the aforementioned nuclear features mention in Cellular Composition Analysis with clinical and pathological features, where it can be seen that both clinical features, age (*p* > 0.05, C-index = 0.59 [95%, 0.59 – 0.60]) and gender (*p* > 0.05, C-index = 0.52 [95%, 0.52 – 0.53]) are nonsignificant. Conversely, the pathological features showed significance for binary grading (*p* = 0.004, C-index = 0.68 [95%, 0.67 – 0.69]) and WHO based grading when moderate and severe cases were combined against mild grade (*p* = 0.04, C-index = 0.68 [95%, 0.67 – 0.68]). When mild and moderate cases were combined and compared against severe, and they showed the same significance as (*p* = 0.04, C-index = 0.68 [95%, 0.67 – 0.68]). The nuclear features extracted from the epithelial layer, basal layer and connective tissue area also showed significance for minimum number of nuclei count (NC) in basal layer (*p* < 0.05, C-index = 0.70 [95%, 0.69 – 0.71]), epithelial layer (*p* < 0.05, C-index = 0.73 [95%, 0.73 – 0.74]) and in PELs (*p* < 0.05, C-index = 0.73 [95%, 0.72 – 0.73]). Figure 6 (B) shows the KM curves for PEL count and (C) epithelium layer NC where it can be seen that both features are statistically significant in differentiating the high risk and low risk lesions with a clear separation between the two groups. Figure 6 (A) shows the hazard ratio (HR) for variation in basal layer NC and epithelium layer NC appears to be associated with improved survival whereas the minimum PEL count, epithelium layer NC and basal layer NC are the adverse predictors of PFS. Furthermore, Table 6 shows the multivariate analysis of most significant nuclear and pathological features (i.e., binary grading, ∧ epithelial layer NC, ∧ basal layer NC and ∧ PEL count) to examine their combined effect on the PFS. When these features are combined, the C-index improves by reaching 0.79 [95%, 0.78 – 0.80] with binary grading, epithelium layer NC and PEL being most significant prognostic features for malignant transformation. In the absence of binary grading, the C-index achieved using nuclear features only is competitive, reaching 0.78 [95%, 0.77 – 0.78]. Similarly, combined binary grading with PEL counts reached the same C-index of 0.78 [95%, 0.77 – 0.78] as compared to the other two feature with binary grading i.e., epithelium layer NC 0.76 [95%, 0.75 – 0.77] and basal layer NC 0.77 [95%, 0.76 – 0.77]. This highlights the importance of using PEL counts as a prognostic feature for predicting the malignant transformation. Further, the combined performance of basal layer NC and epithelium layer NC with PEL count also shows the significance of using PEL in conjunction with other clinical and nuclear features.

**Table 5.** Univariate analysis of the clinical, pathological and digital features where p is calculated using the log-rank method and C-index is calculated using the Cox Proportional Hazard model bootstrapped 1000 times for lower and upper confidence interval.

| Feature | Aggregation | $p$ | C-index | Lower<br>95% | Upper<br>95% |
| --- | --- | --- | --- | --- | --- |
| **Clinical Parameters** |  |  |  |  |  |
| Gender | - | > 0.05 | 0.52 | 0.52 | 0.53 |
| Age | - | > 0.05 | 0.59 | 0.59 | 0.60 |
| **Pathological Parameters** |  |  |  |  |  |
| WHO Grading [Mild vs Mod + Severe] | - | < 0.05 | 0.68 | 0.68 | 0.69 |
| WHO Grading [Mild + Mod vs Severe] | - | < 0.05 | 0.68 | 0.68 | 0.68 |
| Binary Grading | - | < 0.05 | 0.68 | 0.68 | 0.69 |
| **Nuclear Features** |  |  |  |  |  |
| PEL count | $\mu$ | > 0.05 | 0.45 | 0.45 | 0.46 |
| | $\sigma$ | < 0.05 | 0.60 | 0.59 | 0.60 |
| | $m$ | > 0.05 | 0.57 | 0.56 | 0.58 |
| | $\wedge$ | < **0.05** | **0.73** | **0.72** | **0.73** |
| | $\vee$ | > 0.05 | 0.53 | 0.52 | 0.54 |
| Basal NC | $\mu$ | > 0.05 | 0.45 | 0.44 | 0.46 |
| | $\sigma$ | < 0.05 | 0.66 | 0.65 | 0.67 |
| | $m$ | > 0.05 | 0.52 | 0.51 | 0.53 |
| | $\Lambda$ | $< 0.05$ | 0.70 | 0.69 | 0.71 |
| | $\vee$ | $> 0.05$ | 0.53 | 0.52 | 0.54 |
| Epithelium NC | $\mu$ | $< 0.05$ | 0.65 | 0.64 | 0.65 |
| | $\sigma$ | $< 0.05$ | 0.72 | 0.71 | 0.73 |
| | $m$ | $< 0.05$ | 0.66 | 0.65 | 0.67 |
| | $\Lambda$ | **$< 0.05$** | **0.73** | **0.73** | **0.74** |
| | $\vee$ | $> 0.05$ | 0.46 | 0.45 | 0.47 |

**Table 6.** Multivariate analysis of the pathological and digital features where p is calculated using the Wald test and C-index is calculated using the Cox Proportional Hazard model bootstrapped 1000 times for lower and upper confidence interval.

| Feature | $p$ | HR | Lower 95% | Upper 95% |
| --- | --- | --- | --- | --- |
| **C-index = 0.79, 95% CI [0.78 – 0.80]** |  |  |  |  |
| Binary Grading | $< 0.05$ | 2.43 | 1.30 | 4.54 |
| Basal NC | $> 0.05$ | 1.04 | 0.79 | 1.37 |
| PEL count | $< 0.05$ | 1.72 | 1.24 | 2.37 |
| Epithelium NC | $< 0.05$ | 1.48 | 1.07 | 2.05 |
| **C-index = 0.78, 95% CI [0.77 – 0.78]** |  |  |  |  |
| Basal NC | $> 0.05$ | 1.08 | 0.81 | 1.43 |
| PEL count | $< 0.05$ | 1.72 | 1.23 | 2.39 |
| Epithelium NC | $< 0.05$ | 1.67 | 1.20 | 2.32 |
| **C-index = 0.77, 95% CI [0.76 – 0.77]** |  |  |  |  |
| Binary Grading | < 0.05 | 2.97 | 1.62 | 5.43 |
| Basal NC | < 0.05 | 1.54 | 1.31 | 1.81 |
| C-index = 0.78, 95% CI [0.77 – 0.78] |  |  |  |  |
| Binary Grading | < 0.05 | 3.10 | 1.70 | 5.65 |
| PEL count | < 0.05 | 1.81 | 1.50 | 2.18 |
| C-index = 0.76, 95% CI [0.75 – 0.77] |  |  |  |  |
| Binary Grading | < 0.05 | 2.76 | 1.44 | 4.93 |
| Epithelium NC | < 0.05 | 1.84 | 1.27 | 2.66 |
| C-index = 0.73, 95% CI [0.72 – 0.74] |  |  |  |  |
| Basal NC | > 0.05 | 1.13 | 0.84 | 1.52 |
| PEL count | < 0.05 | 1.68 | 1.20 | 2.34 |
| C-index = 0.77, 95% CI [0.77 – 0.78] |  |  |  |  |
| Epithelium NC | < 0.05 | 1.67 | 1.19 | 2.35 |
| Basal layer NC | < 0.05 | 1.54 | 1.29 | 1.83 |
| C-index = 0.78, 95% CI [0.77 – 0.78] |  |  |  |  |
| Epithelium NC | < 0.05 | 1.68 | 1.21 | 2.34 |
| PEL count | < 0.05 | 1.83 | 1.50 | 2.25 |

**Figure 6.**
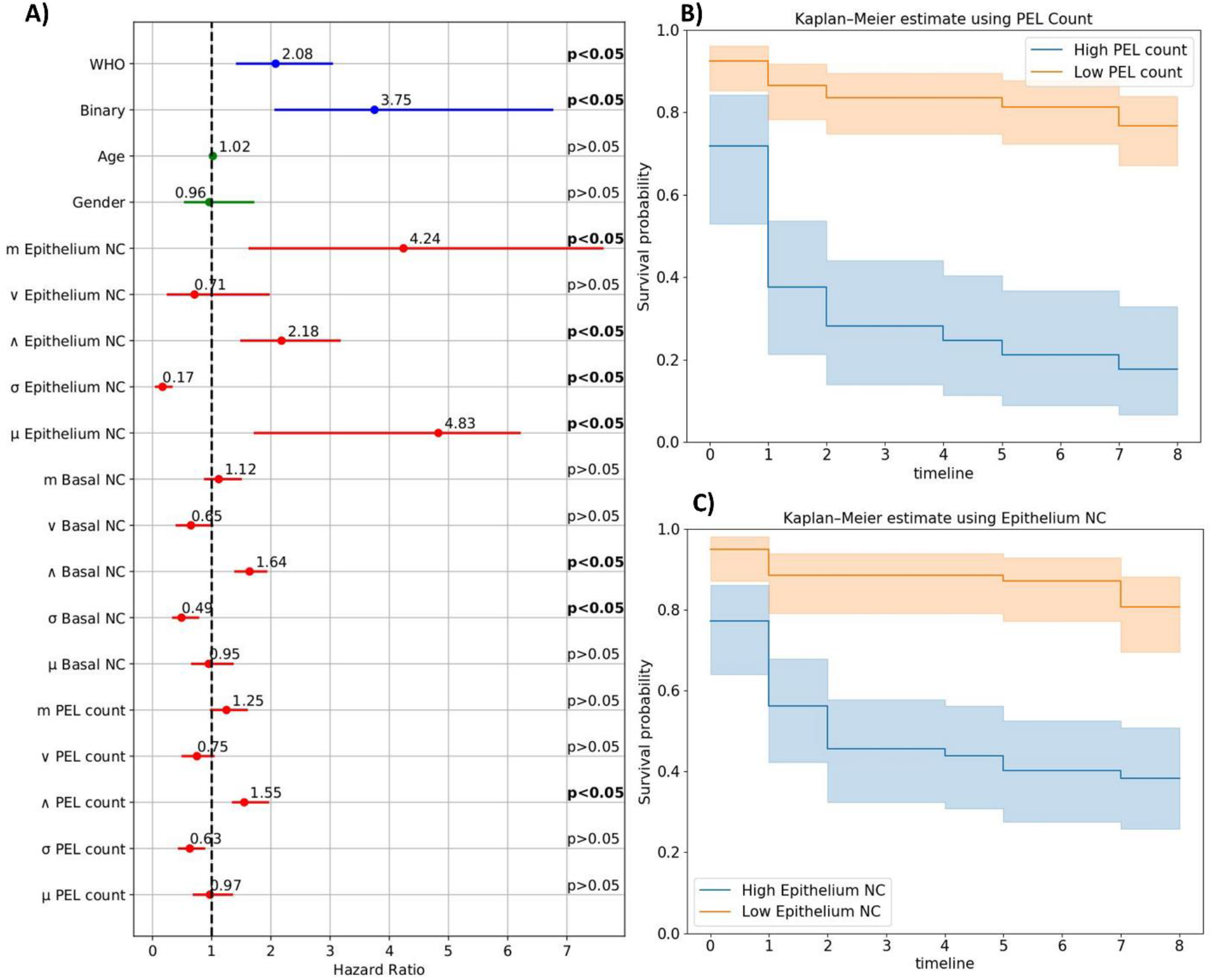
(A) Univariate analysis of different features (blue) pathological (green) clinical and (red) nuclear. For each feature the dot represents the hazards ratio, and the filled line shows the lower and upper confidence interval of 95%. p-values were shown at the right, calculated using the Wald test. (B) Kaplan–Meir (KM) curve for progression free survival of OED using PEL count and (C) represents the KM curve using the epithelium layer nuclei count.

## Discussion and Conclusions

In this study, we explored the potential of deep learning for predicting the malignant transformation from digitised OED histology slides. We trained a weakly supervised learning framework for malignant transformation prediction and further analyse the predictive “hostpots” in epithelial and peri-epithelial tissue regions. We have demonstrated that deep learning based weakly supervised IDaRS can predict malignant transformation with an AUROC of ∼0.78 (±0.07 SD) on stratified 5-fold cross-validation using three different random seeds. Mahmood *et al*. ^32^ also reported the AUROC of 0.77 for transformation using a similar but smaller cohort with the nuclear features subjectively assessed by three pathologists. The higher performance of IDaRS is because it dynamically learns important feature representations from the patches internally, as compared to fixed feature representation of a patch as an input limiting the learning possibilities of the model.

We have also explored the cellular compositions (i.e., nuclear features) and their role in potentially malignant areas (i.e., hotspots) of transformed cases and comparing them to the non-transformed areas (i.e., coldspots). Nuclear features from the epithelial layer and associated connective tissue area were found to be most significant prognostic features for predicting malignant transformation. Other important features found in epithelial and basal layer during the experiments were variation in number of layer nuclei in 100µ per pixel (mpp), standard deviation in cell eccentricity, mean major and minor axis length etc. These nuclear features also correspond to the aberration of nuclei (i.e., variation in size of nuclei captured as a variation in the minor axis of the nuclei and convexity of the nuclear shape) and congestion due proliferation of nuclei in the epithelial and basal layer. However, in order to verify the significance of these features further we require more data to validate these features for their prognostic significance for malignancy. It has also been reported in the literature that the PELs can play an important role in transforming dysplasia into carcinoma ^40^. There is a possible explanation for transformation, that the epithelium is affected by the PEL. This can be due to the release of cytokines linked with oxidative stress, transforming the epithelial cells into premalignant ones ^41–44^ as we have seen that PELs showed significance for predicting the transformation with *p* < 0.05.

For progression free survival, we analysed the clinical, pathological, and nuclear features and found that apart from binary grading in OED, the variation in basal and epithelial layer NC to be associated with improved PFS while minimum number of in basal layer NC, epithelial layer NC and PEL to be associated with increased risk of malignant transformation or poor survival. Gan *et al*. ^40^ has also shown similar finding of PELs using the RNA sequencing of the immune infiltrating sites within the moderate and severe OED. They have stated in their work that the lack of CD8 T-cells in non-cytotoxic subtype and non-immune reactive subtype can lead to progression in moderate and severe dysplasia. It is also interesting to note that our study has found binary grading to be a significant indicator for malignant transformation whereas the study performed by Dost *et al*. ^30^ has shown no association between grading and transformation whereas Mahmood *et al*. ^32^ showed associate between nuclear features and transformation. However, the nuclear features used corresponds to OED grading e.g., bulbus rete pegs, loss of epithelial cohesion etc., and upon adding histological grades into the mix they observed improvements in their results. Similarly, Gilvetti *et al*. ^31^ has shown the significance of different clinical feature specially the age which we have found for one of the sub groups i.e., 0-50 showed prognostic significance with *p* = 0.001. We have also found that in our multivariate analysis that when we combined these pathological and nuclear features for PFS it improved the results specifically due to the addition of epithelium layer NC and PEL count. However, an interesting avenue would be to analyse and investigate the role of dysplasia infiltrating lymphocytes (DILs) in malignant transformation. Although the cohort is small and uni-centric, the department in question is a regional and national referral centre in the UK. Nonetheless, for the practical application and adaptation of these methods in clinical practice requires substantially large and truly multicentric cohort data allowing more rigorous validation of the proposed algorithms.

To best of our knowledge this is the first study to propose and show the role of peri-epithelial lymphocytes (PELs) count in malignant transformation along with other digital biomarkers e.g., epithelium layer NC and basal layer NC. Our multivariate feature analysis has shown that PELs and epithelial NC have shown to improve the prognostic value in conjunction with binary OED grading for predicting malignant transformation. Our proposed methodology for predicting malignancy in an end-to-end manner has the potential play an important role in precision medicine and personalised patient management for early prediction of malignancy risk with the potential to guide treatment decisions and risk stratification.

## Data Availability

All data produced in the present study are available upon reasonable request to the authors

## Acknowledgements

RMSB is funded by the Chancellor Scholarship from University of Warwick. HM is funded by a NIHR Doctoral Research Fellowship. AS, NA, SAK and NMR are funded by a Cancer Research UK Project Grant (ANTICIPATE).

## Statement of author contribution

RMSB, SEAR, SAK and NMR designed the study with help of all co-authors. RMSB, SEAR and NMR developed the computational methods. RMSB wrote the code and carried out all the experiments. SAK and HM obtained the ethical approval, retrieved the histological and clinical data the data. SAK, HM, AS, RMSB and NA provided the WSI annotations. RMSB, SEAR, SAK and NMR were all involved in the drafting of the paper. HM, AS and NA provided peer review for the final draft. All authors read and approved the final paper.

## References

1. WHO. Global Oral Health Status Report. https://www.who.int/news-room/fact-sheets/detail/oral-health. ISBN: 978-92-4-006148-4 (2020).

2. Torre, L. A. et al. Global cancer statistics, 2012. CA. Cancer J. Clin. 65, 87–108 (2015).

3. França, D. C. et al. Unusual Presentation of Oral Squamous Cell Carcinoma in a Young Woman. Sultan Qaboos Univ. Med. J. 12, 228–231 (2012).

4. El-Naggar, A. K., Chan, J. K. C., Grandis, J. R., Takata, T. & Slootweg, P. J. WHO Classification of Head and Neck Tumours. (International Agency for Research on Cancer, 2017).

5. Wright, A. & Shear, M. Epithelial dysplasia immediately adjacent to oral squamous cell carcinomas. J. Oral Pathol. Med. 14, 559–564 (1985).

6. Speight, P. M., Khurram, S. A. & Kujan, O. Oral potentially malignant disorders: risk of progression to malignancy. Oral Surg. Oral Med. Oral Pathol. Oral Radiol. 125, 612–627 (2018).

7. Kujan, O. et al. Evaluation of a new binary system of grading oral epithelial dysplasia for prediction of malignant transformation. Oral Oncol. 42, 987–993 (2006).

8. Kujan, O. et al. Why oral histopathology suffers inter-observer variability on grading oral epithelial dysplasia: An attempt to understand the sources of variation. Oral Oncol. 43, 224– 231 (2007).

9. Shephard, A. J. et al. Simultaneous Nuclear Instance and Layer Segmentation in Oral Epithelial Dysplasia. in 552–561 (2021).

10. Shaban, M. et al. Context-Aware Convolutional Neural Network for Grading of Colorectal Cancer Histology Images. IEEE Trans. Med. Imaging 39, 2395–2405 (2020).

11. Qaiser, T. & Rajpoot, N. M. Learning Where to See: A Novel Attention Model for Automated Immunohistochemical Scoring. IEEE Trans. Med. Imaging 38, 2620–2631 (2019).

12. Simon Graham et al. Classification of lung cancer histology images using patch-level summary statistics. in vol. 10581 (2018).

13. Bejnordi, B. E. et al. Diagnostic Assessment of Deep Learning Algorithms for Detection of Lymph Node Metastases in Women With Breast Cancer. JAMA 318, 2199–2210 (2017).

14. Bashir, R. M. S. et al. Automated grade classification of oral epithelial dysplasia using morphometric analysis of histology images. in Medical Imaging 2020: Digital Pathology vol. 11320 1132011 (International Society for Optics and Photonics, 2020).

15. Graham, S. et al. Hover-Net: Simultaneous segmentation and classification of nuclei in multi-tissue histology images. Med. Image Anal. 58, 101563 (2019).

16. Sirinukunwattana, K. et al. Locality Sensitive Deep Learning for Detection and Classification of Nuclei in Routine Colon Cancer Histology Images. IEEE Trans. Med. Imaging 35, 1196–1206 (2016).

17. Akram, S. U. et al. Leveraging Unlabeled Whole-Slide-Images for Mitosis Detection. in Computational Pathology and Ophthalmic Medical Image Analysis (eds. Stoyanov, D.et al.) 69–77 (Springer International Publishing, 2018). doi:10.1007/978-3-030-00949-6_9.

18. Qaiser, T. et al. Digital Tumor-Collagen Proximity Signature Predicts Survival in Diffuse Large B-Cell Lymphoma. in Digital Pathology (eds. Reyes-Aldasoro, C. C., Janowczyk, A., Veta, M., Bankhead, P. & Sirinukunwattana, K.) 163–171 (Springer International Publishing, 2019). doi:10.1007/978-3-030-23937-4_19.

19. Graham, S. et al. MILD-Net: Minimal information loss dilated network for gland instance segmentation in colon histology images. Med. Image Anal. 52, 199–211 (2019).

20. Fraz, M. M. et al. FABnet: feature attention-based network for simultaneous segmentation of microvessels and nerves in routine histology images of oral cancer. Neural Comput. Appl. 32, 9915–9928 (2020).

21. Gamper, J. et al. PanNuke Dataset Extension, Insights and Baselines. ArXiv200310778 Cs Eess Q-Bio (2020).

22. Shaban, M. et al. A Novel Digital Score for Abundance of Tumour Infiltrating Lymphocytes Predicts Disease Free Survival in Oral Squamous Cell Carcinoma. Sci. Rep. 9, 13341 (2019).

23. Sami, M. M., Saito, M., Muramatsu, S., Kikuchi, H. & Saku, T. A Computer-Aided Distinction Method of Borderline Grades of Oral Cancer. IEICE Trans. Fundam. Electron. Commun. Comput. Sci. E93.A, 1544–1552 (2010).

24. Nag, R., Chatterjee, J., Paul, R. R., Pal, M. & Das, R. K. Nuclear Segmentation and its Quantification in H E Stained Images of Oral Precancer to Detect its Malignant Potentiality. in 2018 International Conference on Current Trends towards Converging Technologies (ICCTCT) 1–6 (2018). doi:10.1109/ICCTCT.2018.8550984.

25. Adel, D. et al. Oral Epithelial Dysplasia Computer Aided Diagnostic Approach. in 2018 13th International Conference on Computer Engineering and Systems (ICCES) 313–318 (2018). doi:10.1109/ICCES.2018.8639452.

26. Das, D. K. et al. Automatic identification of clinically relevant regions from oral tissue histological images for oral squamous cell carcinoma diagnosis. Tissue Cell 53, 111–119 (2018).

27. Das, N., Hussain, E. & Mahanta, L. B. Automated classification of cells into multiple classes in epithelial tissue of oral squamous cell carcinoma using transfer learning and convolutional neural network. Neural Netw. 128, 47–60 (2020).

28. Silva, A. B. et al. Computational analysis of histological images from hematoxylin and eosin-stained oral epithelial dysplasia tissue sections. Expert Syst. Appl. 193, 116456 (2022).

29. Nguyen, P.-T.-H., Sakamoto, K. & Ikeda, T. Deep-learning application for identifying histological features of epithelial dysplasia of tongue. J. Oral Maxillofac. Surg. Med. Pathol. (2022) doi:10.1016/j.ajoms.2021.12.008.

30. Dost, F., Lê Cao, K., Ford, P. J., Ades, C. & Farah, C. S. Malignant transformation of oral epithelial dysplasia: a real-world evaluation of histopathologic grading. Oral Surg. Oral Med. Oral Pathol. Oral Radiol. 117, 343–352 (2014).

31. Gilvetti, C., Soneji, C., Bisase, B. & Barrett, A. W. Recurrence and malignant transformation rates of high grade oral epithelial dysplasia over a 10 year follow up period and the influence of surgical intervention, size of excision biopsy and marginal clearance in a UK regional maxillofacial surgery unit. Oral Oncol. 121, 105462 (2021).

32. Mahmood, H. et al. Prediction of malignant transformation and recurrence of oral epithelial dysplasia using architectural and cytological feature specific prognostic models. Mod. Pathol. 1–9 (2022) doi:10.1038/s41379-022-01067-x.

33. He, K., Zhang, X., Ren, S. & Sun, J. Deep Residual Learning for Image Recognition. in 2016 IEEE Conference on Computer Vision and Pattern Recognition (CVPR) 770–778 (2016). doi:10.1109/CVPR.2016.90.

34. Bilal, M. et al. Development and validation of a weakly supervised deep learning framework to predict the status of molecular pathways and key mutations in colorectal cancer from routine histology images: a retrospective study. Lancet Digit. Health 3, e763–e772 (2021).

35. Ilse, M., Tomczak, J. & Welling, M. Attention-based Deep Multiple Instance Learning. in Proceedings of the 35th International Conference on Machine Learning 2127–2136 (PMLR, 2018).

36. Lu, M. Y. et al. Data Efficient and Weakly Supervised Computational Pathology on Whole Slide Images. ArXiv200409666 Cs Eess Q-Bio (2020).

37. Huang, G., Liu, Z., van der Maaten, L. & Weinberger, K. Q. Densely Connected Convolutional Networks. ArXiv160806993 Cs (2018).

38. Dosovitskiy, A. et al. An Image is Worth 16×16 Words: Transformers for Image Recognition at Scale. ArXiv201011929 Cs (2021).

39. Benjamini, Y. & Hochberg, Y. Controlling the False Discovery Rate: A Practical and Powerful Approach to Multiple Testing. J. R. Stat. Soc. Ser. B Methodol. 57, 289–300 (1995).

40. Gan, C. P. et al. Transcriptional analysis highlights three distinct immune profiles of high-risk oral epithelial dysplasia. Front. Immunol. 13, 954567 (2022).

41. Fitzpatrick, S. G., Honda, K. S., Sattar, A. & Hirsch, S. A. Histologic lichenoid features in oral dysplasia and squamous cell carcinoma. Oral Surg. Oral Med. Oral Pathol. Oral Radiol. 117, 511–520 (2014).

42. Grivennikov, S. I., Greten, F. R. & Karin, M. Immunity, Inflammation, and Cancer. Cell 140, 883–899 (2010).

43. Rhodus, N. L. et al. The feasibility of monitoring NF-κB associated cytokines: TNF-α, IL-1α, IL-6, and IL-8 in whole saliva for the malignant transformation of oral lichen planus. Mol. Carcinog. 44, 77–82 (2005).

44. Georgakopoulou, E. A., Achtari, M. D., Achtaris, M., Foukas, P. G. & Kotsinas, A. Oral Lichen Planus as a Preneoplastic Inflammatory Model. BioMed Res. Int. 2012, e759626 (2012).

